# Cardiovascular Complications with Delivery Hospitalizations in Patients with Pulmonary Hypertension: A Nationwide Study from 2011-2020

**DOI:** 10.1101/2023.07.06.23292327

**Authors:** Ankit Agrawal, Suryansh Bajaj, Umesh Bhagat, Sanya Chandna, Aro Daniela Arockiam, Yasser Jamil, Mahmoud El Iskandarani, Rahul Gupta, Divya Nayar, Erin D. Michos

**Affiliations:** Department of Cardiovascular Medicine, Heart, Vascular, and Thoracic Institute, Cleveland Clinic, Cleveland, Ohio; Department of Radiology, University of Arkansas for Medical Sciences, Little Rock, Arkansas; Department of Hospital Medicine, Cleveland Clinic, Cleveland, Ohio; Department of Internal Medicine, Yale School of Medicine, New Haven, Connecticut; Department of Internal Medicine, Eastern Connecticut Health Network, Manchester, Connecticut; Lehigh Valley Heart Institute, Lehigh Valley Health Network, Allentown, Pennsylvania; Department of Neurology, University of Arkansas for Medical Sciences, Little Rock, Arkansas; Division of Cardiology, Johns Hopkins University, School of Medicine, Baltimore, MD

**Keywords:** Hypertension, Pulmonary, Pregnancy, Congestive heart failure, Cardiac arrhythmias

## Abstract

**Background:** Pregnancy in patients with pulmonary hypertension (PH) is associated with heightened risk of various medical complications. Our study aims to understand the patient characteristics and investigate the association between PH and these complications in pregnant patients during delivery.

**Methods:** The National Inpatient Sample (NIS) was used to identify delivery hospitalizations from 2011 to 2020. The primary outcomes were in-hospital medical and obstetric complications. Multivariate logistic regression was performed to study the association of PH with these complications.

**Results:** A total of 37,482,207 delivery hospitalizations in women ≥18 years were identified from the NIS database out of which 9,593 patients had PH. Pregnant patients with PH had a higher incidence of complications during delivery including preeclampsia/eclampsia, cardiac arrhythmias, pulmonary edema amongst others, compared to pregnant patients without PH. Pregnant patients with PH had a higher incidence of in-hospital mortality compared to those without PH (0.51% vs 0.007%). In adjusted analyses, PH was independently associated with a higher risk of pulmonary edema (OR: 18.65 [95% CI: 13.71-25.38]), peripartum cardiomyopathy (14.06 [9.15-21.60]), venous thromboembolism (12.25 [7.80-19.24]), cardiac arrhythmias (11.75 [10.11-13.67]), acute kidney injury (7.53 [5.36-10.58]), preeclampsia/eclampsia (4.61 [4.04-5.25]), and acute coronary syndrome (2.83 [1.17-6.85]), compared with pregnant patients without PH. In-hospital mortality in patients with PH was associated with stroke (127.33 [78.49-206.57]), acute kidney injury (51.25 [34.40-76.36]), cardiac arrhythmias (24.80 [19.43-31.65]), peripartum cardiomyopathy (6.47 [3.23-12.97]), pulmonary edema (4.27 [2.18-8.37]), venous thromboembolism (2.75 [1.07-7.10]), and preeclampsia/eclampsia (1.87 [1.35-2.60]) compared to pregnant patients without PH.

**Conclusion:** Delivery hospitalizations in patients with PH are associated with high risk of various complications. Prenatal counseling and multidisciplinary care are essential to help mitigate unfavorable outcomes in these patients.

## Introduction

Pulmonary hypertension (PH) is a condition characterized by abnormally elevated pressures in the pulmonary vasculature (>25 mmHg) causing right heart strain which may lead to right heart failure. It may be isolated or occur secondary to cardiac, respiratory, or other diseases.^1, 2^ Pregnancy in patients with PH is associated with high risk of maternal, obstetric, and fetal complications and patient mortality.^3–5^ The hemodynamic changes during pregnancy, labor, and postpartum period lead to increased cardiac output and decreased systemic vascular resistance that can lead to a further increase in pulmonary vascular resistance.^6^ Pregnancy is often considered to be contraindicated in PH patients; however, recent data show that the prevalence of pregnancy in patients with PH is increasing in the United States (US) despite the associated complications.^7–9^ While termination of pregnancy is advised, many patients opt to continue the pregnancy given the advances in management. This underscores the need to better understand the complications of pregnancy in PH patients. A prior analysis using older US data from 2003-2012 previously described the associated increased risk of cardiovascular events in pregnant persons with PH at delivery.^4^ Using more recent data from the years 2011-2020 [notably including the pandemic year of 2020], this nationwide study aimed to evaluate patient characteristics of pregnant persons with PH undergoing delivery, the prevalence of maternal complications at delivery, and the association of PH with these complications.

## Methods

### Data source

The National Inpatient Sample (NIS) database from 2011 to 2020 was used to conduct this study. The NIS is the largest inpatient healthcare database managed by the Agency for Healthcare Research and Quality via Healthcare Cost and Utilization Project (HCUP). About 20% hospitals of all the states participating in HCUP contribute to NIS which contains data of >7 million hospitalization annually that is representative of >97% of the US population. State, hospital, and patient identifiers are completely absent from the database to ensure patient confidentiality. Institutional Review Board approval is not required for this study as NIS uses publicly available anonymous and de-identified data.

### Study Population and covariates

International Classification of Diseases, Tenth Revision, Clinical modification (ICD-10-CM) codes were used to identify delivery hospitalizations. In the cohort, ICD-9 codes 415.0, 416.0, 416.1, 416.8, 416.9, 417.8, 417.9 and ICD-10 codes I27.0, I27.2, I27.8, I27.9, I28.0, I28.8, I28.9 were used to identify patients previously diagnosed with PH. Individuals with age <18 years were excluded from our study. Variables including demographic characteristics (age, race, region, median income, insurance) and clinical history (hypertension, diabetes mellitus, hyperlipidemia, obesity, chronic heart failure, coronary artery disease, chronic kidney disease, and smoking status) were collected for analysis. Maternal complications at delivery were compared between pregnant patients with PH versus those without PH. The primary outcomes of interest were pre-eclampsia, eclampsia, peripartum cardiomyopathy (PPCM), acute heart failure, acute coronary syndrome, ischemic and hemorrhagic stroke, pulmonary edema, cardiac arrhythmias, acute kidney injury (AKI), and venous thromboembolism. Given small numbers of eclampsia, these cases were combined with preeclampsia. In secondary analysis we also examined which maternal complications were associated with in-hospital mortality.

### Statistical Analysis

Descriptive statistics are presented as mean ± standard deviation for continuous variables and percentages for categorical variables. Chi-square test or Fisher’s exact test as appropriate for categorical variables and the Mann–Whitney U test for continuous variables to assess statistical significance. Cochran–Mantel–Haenszel test was performed to derive unadjusted odds ratios (uOR). Multivariate logistic regression model adjusted for age, race or ethnicity, region, chronic hypertension, dyslipidemias, heart failure, coronary artery disease, chronic kidney disease, obesity, smoking, multiple gestation, cesarean delivery, insurance, and median household income was performed. A p-value of <0.05 was considered statistically significant, and all tests were two-tailed. Forest plots were created using Microsoft Excel (Redmond, WA, USA). The analysis was conducted using StataCorp 2021 [Stata Statistical Software: Release 17. StataCorp LLC (College station, TX, USA)]. Appropriate weights provided by HCUP to generate national estimates were used in all estimations.

## Results

### Study population

A total of 37,482,207 delivery hospitalizations in pregnant patients with age ≥18 years were identified from the NIS database from 2011 to 2020, out of which 9,593 patients (26 per 100,000 delivery hospitalization) were previously diagnosed with PH. Compared to delivery hospitalization in patients without PH, those with PH were slightly older (30.6% vs. 28.7%), more likely to have multiple gestations (4.4% vs. 2%) and history of smoking (9.8% vs. 5.3%) and less likely to have a cesarean delivery (40.2% vs. 47.2%). The prevalence of PH was higher in Black patients (32.8% of patients with PH vs.14.8% of patients without PH) than White patients (36% vs. 52.6%), and patients with PH were more likely to be insured by Medicaid (56.1% vs. 42.5%) than private insurance (34.8% vs. 51.3%). Congenital heart disease and autoimmune conditions (systemic lupus erythematosus, systemic sclerosis, sarcoidosis) constituted 2.4% and 4.1%, respectively, as etiologies of PH. Amongst comorbidities, patients with PH were also more likely to have hypertension (8.7% vs. 0.4%), diabetes mellitus (7.2% vs. 1.1%), hyperlipidemia (1.8% vs. 0.2%), obesity (26.6% vs. 8.9%), chronic kidney disease (2.8% vs. 0.09%),chronic heart failure (8.4% vs. 0.03%) and coronary artery disease (2.3% vs.0.03%). The detailed baseline characters of the study population are described in **Table 1**.

**Table 1.**
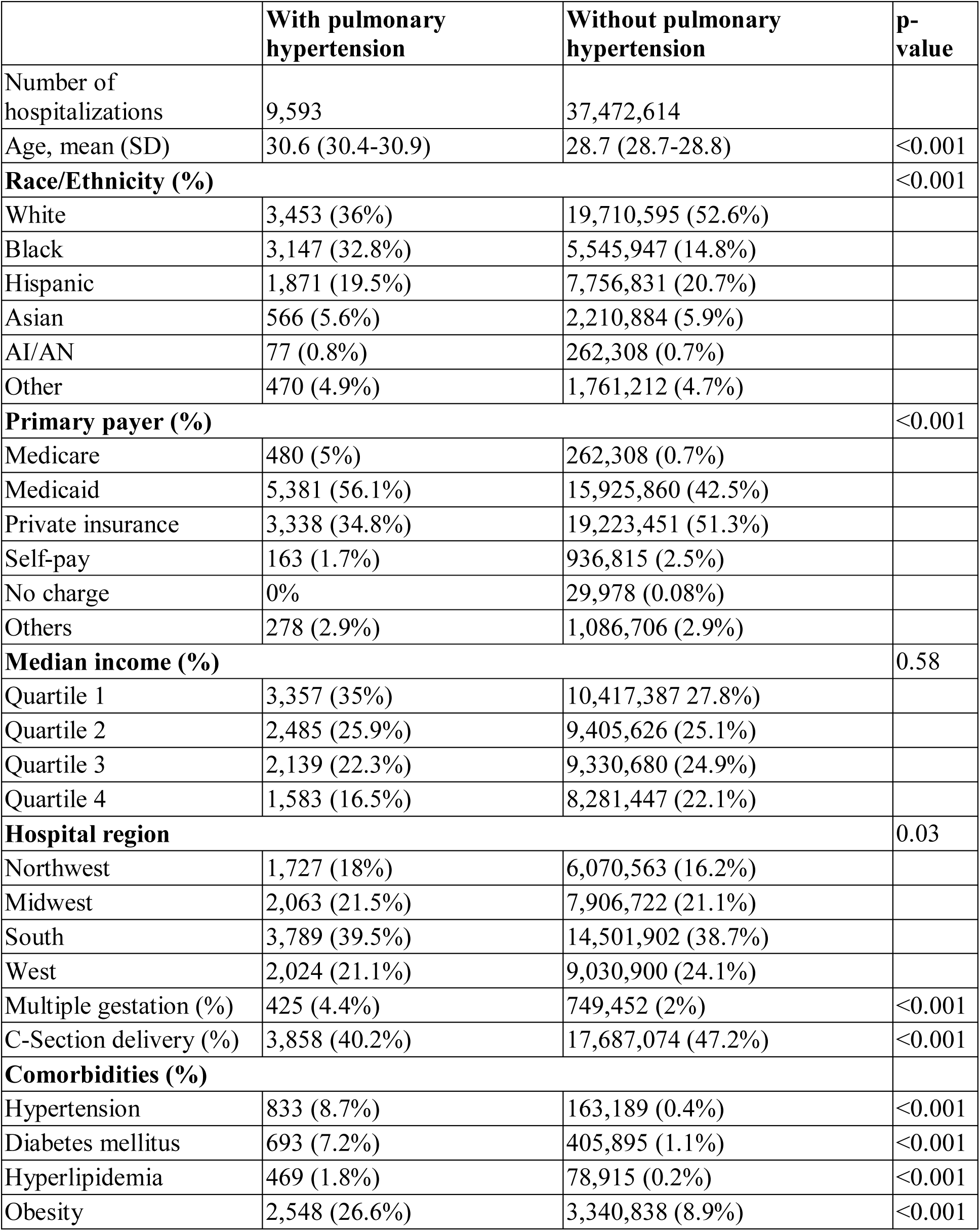
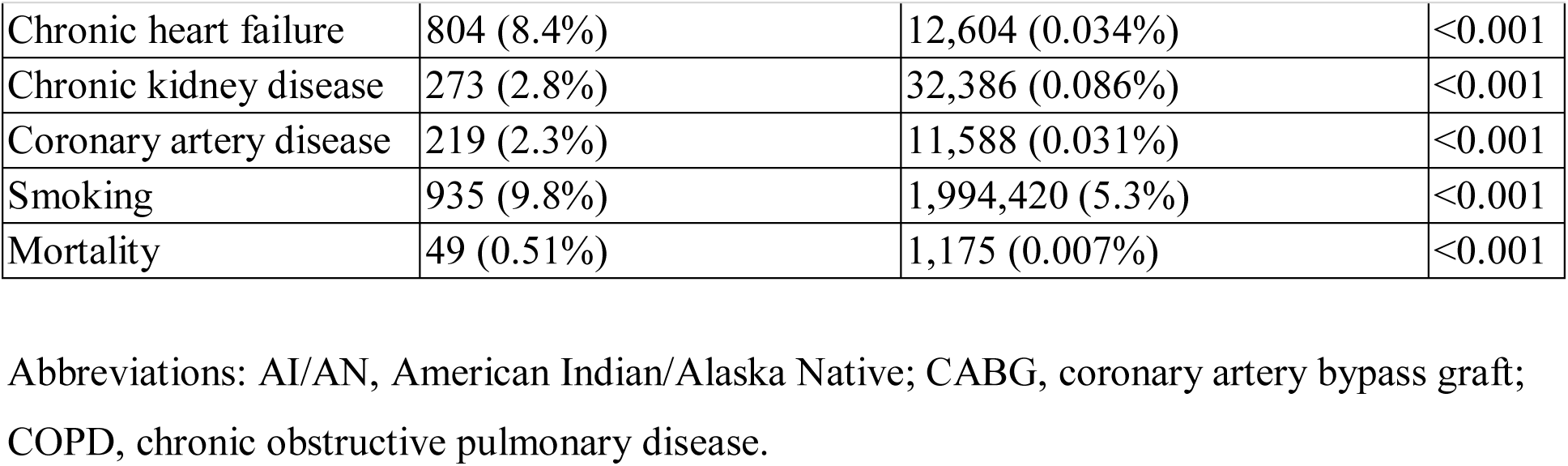
Baseline characteristics of delivery hospitalizations with and without pulmonary hypertension

### Trend of delivery hospitalizations in patients with PH over 10 years

On trend analysis, the prevalence of delivery hospitalizations in patients with PH ranged from 0.022% to 0.03%, with a total average of 0.026% for the entire decade (**Figure 1**).

**Figure 1:**
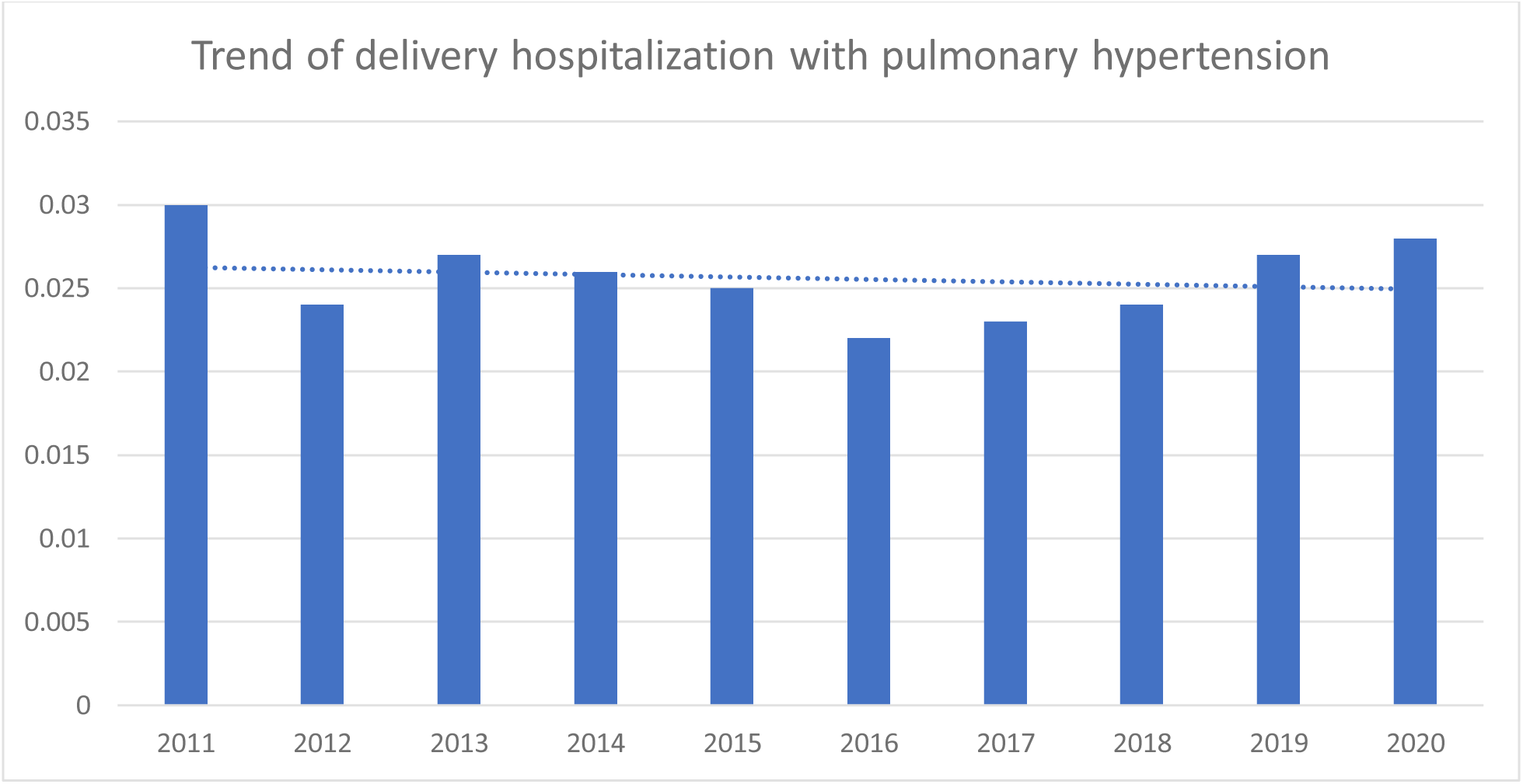
Trend of US delivery hospitalization with pulmonary hypertension as percentages of total annual delivery hospitalization from 2011-2020

### Incidence of in-hospital complications

Pregnant patients with PH had higher rates (per 100,00 delivery hospitalizations) of PPCM (3,690 vs 18), AKI (5,087 vs 82), acute coronary syndrome (813 vs 7), stroke (313vs 19), pulmonary edema (6,044 vs 49), cardiac arrhythmias (15,647 vs931), venous thromboembolism (1,970 vs 55), and preeclampsia/eclampsia (19,222 vs 3,941) compared to pregnant patients without PH. Patients with PH also had a longer length of stay (mean = 6.9 days) than those without PH (mean = 2.6 days). Incidence of in-hospital complications is described in **Table 2**.

**Table 2.** Complication Rates (per 100,000 Delivery Hospitalizations) and Hospital Resource Use in Patients with and without pulmonary hypertension

| **Variables** | **With pulmonary hypertension** | **Without pulmonary hypertension** | **P value** |
| --- | --- | --- | --- |
| **Complication rates (per 100,000 delivery hospitalizations)** |  |  |  |
| Peripartum cardiomyopathy | 3,690 | 18 | <0.01 |
| Acute heart failure | <11* | <11* |  |
| Acute kidney injury | 5,087 | 82 | <0.01 |
| Acute coronary syndrome | 813 | 7 | <0.01 |
| Stroke | 313 | 19 | <0.01 |
| Pulmonary edema | 6,004 | 49 | <0.01 |
| Cardiac arrhythmias | 15,647 | 931 | <0.01 |
| Venous thromboembolism | 1,970 | 55 | <0.01 |
| Preeclampsia/eclampsia | 19,222 | 3,941 | <0.01 |
| **Hospital resource utilization** |  |  |  |
| Mean (IQR) length of stay, days | 6.9 (6.5-7.4) | 2.6 (2.6-2.7) | <0.01 |

\*Cases less than 11 cannot be reported per HCUP guidelines

### Association of PH with in-hospital complications

Multivariable logistic regression model showed that PH at delivery hospitalization was independently associated with a higher risk of pulmonary edema (OR: 18.65[95%CI: 13.71-25.38], p<0.001), PPCM (OR: 14.06 [95%CI: 9.15-21.60], p<0.001), venous thromboembolism (OR: 12.25 [95%CI: 7.80-19.24], p<0.001),cardiac arrhythmias (OR: 11.75[95%CI: 10.11-13.67], p<0.001),AKI (OR: 7.53[95%CI: 5.36-10.58], p<0.001), preeclampsia/eclampsia (OR: 4.61 [95%CI 4.04-5.25], p<0.001), and acute coronary syndrome (OR: 2.83 [95%CI: 1.17-6.85], p=0.022), compared with pregnant patients without PH. The unadjusted analysis showed an increased risk of stroke (OR: 16.40[95%CI: 7.32-36.75], p<0.001) in patients with PH; however, this association was not significant on multivariable analysis (p=0.554). These associations are shown in **Table 3**. A separate sensitivity analysis was performed for the pandemic year 2020 to assess the impact of COVID-19. Multivariate analysis showed that PH was still strongly associated with the occurrence of in-hospital PPCM (OR: 31.90 [11.74-86.69], p<0.001) and pulmonary edema (OR: 9.96 [3.78-26.23], p<0.001) compared to pregnant patients without PH. Further analysis is available in the supplementary analysis (**Supplemental Table 1**).

**Table 3.** Adjusted and unadjusted odds ratio for in-hospital complications among patients with pulmonary hypertension versus without pulmonary hypertension.

|  | **Pulmonary hypertension** |  |
| --- | --- | --- |
| **Complications** | **Odds Ratio (95% CI)** | **P-Value** |
| **Unadjusted analysis (uOR)** |  |  |
| Preeclampsia/Eclampsia | 6.58 (5.86-7.38) | <0.001 |
| Peripartum cardiomyopathy | 207.10 (162.36-264.16) | <0.001 |
| Acute heart failure | 1 (omitted) |  |
| Acute kidney injury | 65.20 (52.81-80.51) | <0.001 |
| Acute coronary syndrome | 118.45 (71.85-195.28) | <0.001 |
| Stroke | 16.40 (7.32-36.75) | <0.001 |
| Pulmonary edema | 130.08 (107.35-157.62) | <0.001 |
| Cardiac arrhythmia | 19.73 (17.46-22.30) | <0.001 |
| Venous thromboembolism | 36.70 (26.60-50.65) | <0.001 |
| **Multi-Variable Analysis (aOR)** |  |  |
| Preeclampsia/Eclampsia | 4.61 (4.04-5.25) | <0.001 |
| Peripartum cardiomyopathy | 14.06 (9.15-21.60) | <0.001 |
| Acute heart failure | 1 (omitted) |  |
| Acute kidney injury | 7.53 (5.36-10.58) | <0.001 |
| Acute coronary syndrome | 2.83 (1.17-6.85) | 0.022 |
| Stroke | 1.38 (0.47-4.13) | 0.554 |
| Pulmonary edema | 18.65 (13.71-25.38) | <0.001 |
| Cardiac arrhythmia | 11.75 (10.11-13.67) | <0.001 |
| Venous thromboembolism | 12.25 (7.80-19.24) | <0.001 |

aOR indicates adjusted odds ratio; and uOR, unadjusted odds ratio. Each condition was adjusted for all the other mentioned variables in the table for multivariate analysis.

### Association of PH with in-hospital mortality

Multivariable analysis showed that important factors for in-hospital mortality in pregnant patients with PH include: stroke (OR: 127.33 [95%CI 78.49-206.57], p<0.001), AKI (OR: 51.25 [95%CI 34.40-76.36], p<0.001), cardiac arrhythmias (OR: 24.80 [95%CI 19.43-31.65], p<0.001), PPCM (OR: 6.47 [95%CI 3.23-12.97], p<0.001), pulmonary edema (OR: 4.27 [95%CI 2.18-8.37], p<0.001), venous thromboembolism (OR: 2.75 [95%CI 1.07-7.10], p=0.036), and preeclampsia/eclampsia (OR: 1.87 [95%CI 1.35-2.60], p<0.001) compared to pregnant patients without PH. The unadjusted analysis showed a strong association of acute coronary syndrome (OR: 373.75 [95%CI 218.20-640.17], p<0.001) with in-hospital mortality in PH patients; however, this association was not significant on multivariable analysis (p=0.067). These associations are shown in **Table 4**.A separate sensitivity analysis was performed for the year 2020 to assess the impact of COVID-19. Multivariate analysis showed that mortality in pregnant patients with PH was still strongly associated with AKI (OR: 126.29 [44.35-359.58], p<0.001) and stroke (OR: 29.89 [1.77-504.38], p=0.01) compared to pregnant patients without PH. Further analysis is available in the supplementary analysis (**Supplemental Table 2**).

**Table 4:** Adjusted and unadjusted odds ratio for complications associated with in-hospital patient mortality among patients with pulmonary hypertension versus without pulmonary hypertension.

|  | **Patient mortality** |  |
| --- | --- | --- |
| **Complications** | **Odds Ratio (95% CI)** | **P-Value** |
| **Unadjusted analysis (uOR)** |  |  |
| Preeclampsia/Eclampsia | 5.79 (4.63-7.24) | <0.001 |
| Peripartum cardiomyopathy | 299.35 (206.21-434.58) | <0.001 |
| Acute heart failure | 1 (omitted) |  |
| Acute kidney injury | 342.56 (278.84-420.83) | <0.001 |
| Acute coronary syndrome | 373.75 (218.20-640.17) | <0.001 |
| Stroke | 684.86 (520.40-901.30) | <0.001 |
| Pulmonary edema | 101.42 (68.94-149.20) | <0.001 |
| Cardiac arrhythmia | 47.08 (38.98-56.87) | <0.001 |
| Venous thromboembolism | 57.68 (35.62-93.39) | <0.001 |
| **Multi-Variable Analysis (aOR)** |  |  |
| Preeclampsia/Eclampsia | 1.87 (1.35-2.60) | <0.001 |
| Peripartum cardiomyopathy | 6.47 (3.23-12.97) | <0.001 |
| Acute heart failure | 1 (omitted) |  |
| Acute kidney injury | 51.25 (34.40-76.36) | <0.001 |
| Acute coronary syndrome | 2.84 (0.93-8.66) | 0.067 |
| Stroke | 127.33 (78.49-206.57) | <0.001 |
| Pulmonary edema | 4.27 (2.18-8.37) | <0.001 |
| Cardiac arrhythmia | 24.80 (19.43-31.65) | <0.001 |
| Venous thromboembolism | 2.75 (1.07-7.10) | 0.036 |

aOR indicates adjusted odds ratio; and uOR, unadjusted odds ratio. Each condition was adjusted for all the other mentioned variables in the table for multivariate analysis.

## Discussion

Our US nationwide study from 2011 to 2020 found that PH is associated with various maternal complications at the time of delivery hospitalizations compared to pregnant patients without PH. While the overall prevalence of PH is low (26 per 100,000 delivery hospitalizations), previous studies have shown that the prevalence of PH in pregnant patients has been increasing with these patients making up 6.5% of all the pregnant patients with heart disease.^8^ Patients with PH were found to be more likely to be of Black race/ethnicity, have other comorbidities, and were twice more likely to have multiple gestation, although the rates of cesarean section deliveries were lower compared to delivery hospitalizations in patients without PH. In line with previous studies, preeclampsia/eclampsia and cardiac arrhythmias were the most commonly seen complications followed by pulmonary edema, AKI, and PPCM.^3, 4^ PH was found to be strongly associated with cardiac complications including pulmonary edema, cardiac arrhythmias, and embolic phenomena amongst other conditions. These complications led to higher in-hospital mortality incidence (0.51% vs 0.007%) and more than double the length of hospital stay (6.9 days) compared to patients without PH (2.6 days), in turn leading to higher expenditure. A prior study showed that the total hospital charges for patients with PH (mean = $41,171) were more than thrice compared to those without PH (mean = $12,036) and over 50% of these patients are insured by Medicaid. This creates an additional burden on patients as well as the hospital resources.^4^

During the pandemic year 2020, delays or avoidance of medical care were reported due to fears of contracting COVID-19;^10^ therefore, in sensitivity analysis, we examined outcomes from this year separately. In our subgroup analysis from the year 2020, the most important in-hospital complication associated with pregnant patients with PH was PPCM. Further, the most important factor associated with in-hospital mortality was AKI. This difference from the overall results can stem from the multi-system disease caused by COVID-19 which potentially adds to the existing co-morbidities, or perhaps to the aforementioned delays in medical care. Further large-scale study needs to be performed to identify the impact of COVID-19 on in-hospital outcomes for pregnant patients with PH, specifically. However, the impact of COVID-19 on in-hospital outcomes for pregnant patients in the overall NIS cohort has recently been reported.^11^

The European Society of Cardiology (ESC) and the European Respiratory Society (ERS) guidelines recommend avoiding pregnancy or early termination of pregnancy in patients with PH due to the associated cardiovascular risks to the patients.^2, 12^ With advances in medicine and improvement in access to care, the mortality rates in patients with PH have drastically reduced from more than 50% to 0.8%-3.3%, with one study reporting zero maternal deaths in PH patients during pregnancy and delivery.^3–5, 13, 14^ Our study found the in-hospital mortality rate of pregnant patients with PH to be 0.51%. However, it has also been pointed out that studies with lower mortality rates often lack follow-up data on the patient cohort which might lead to underestimation of true mortality rates,^15^ and the highest risk patients with PH (such as those with Eisenmenger’s physiology) may have avoided pregnancy altogether as per guideline recommendations. Apart from the prepartum period and the labor, the postpartum period up to 6 months post-delivery is also considered to be high risk for these patients due to the ongoing hemodynamic fluctuations. These fluctuations may lead to additional complications and even maternal mortality within first year after delivery.^12^ Moreover, some of the patients are not conclusively diagnosed with PH by right heart catheterization but instead are only diagnosed based on the echocardiography findings which may have led to over-diagnosis of true PH and thus inclusion of patients with more favorable outcomes. Nevertheless, the high rates of morbidity associated with the complications demonstrate that significant risks of pregnancy in patients with PH remain. Our study also showed that patients with PH have 2.5-3 times higher risk of developing maternal complications like preeclampsia/eclampsia, and 1.3 times higher risk of peripartum cardiomyopathy. Patients with PH are also known to obstetric bleeding, postpartum hemorrhage, and postpartum infection.^4^ Additionally, patients with PH also have higher risk of adverse fetal outcomes. Sliwa et al showed that neonatal death rate was 9.3% up to 1 week of life and 20% of the neonates born to patients with PH were small for gestation age (<2,500 g) with 4% having some cardiac anomaly.^3^

In pregnant patients with group 1 PH (pulmonary arterial hypertension, PAH), PAH associated with other systemic or connective tissue disease have higher mortality rates compared to PAH associated with congenital heart disease and idiopathic PAH.^13^ Patients with PAH secondary to other diseases or due to congenital heart disease have higher risk of having low birth weight neonates compared to those with idiopathic PAH. Patients with group 1 PH also more commonly undergo cesarean section deliveries as compared to group 2 PH (PH due to heart disease).^3^ Patients with group 2 PH (PH due to heart disease) have higher risk of cardiac complications (heart failure and cardiac arrhythmias), obstetric complications (preeclampsia and eclampsia) and the risk of preterm birth compared to patients with group 1 PH, although the rate of cesarean section deliveries is higher for patients with group 1 PH.^3, 4^ The mode of delivery for PH patients is debatable. While the vaginal delivery is associated with hemodynamic changes during uterine contractions at the time of labor, cesarean section has complications related to risk of blood loss, infection, potential hypotensive effects if regional anesthesia used, and increase in pulmonary vascular resistance due to positive pressure ventilation and decreased cardiac index if general anesthesia required.^6, 16^ The common consensus is that patients with PH should undergo cesarean section deliveries; however, our study showed that it is less commonly performed in delivery hospitalizations in patients with PH compared to other patients.^4, 6^ Nevertheless, a multidisciplinary cardio-obstetrics team is needed to provide optimal care to these patients owing to the risk of complications as evidenced by our study.^17, 18^

## Limitations

Our study has a few limitations. Firstly, we used the NIS database to extract the patient cohort. While the NIS provides a lot of useful data, information about severity of PH is not present. Secondly, NIS only collects the inpatient data for delivery hospitalizations and does not provide information on the follow up of these patients especially during the high-risk postpartum period. Lastly, there is a lack of information on lab value and medication history of patients which might help better understand the association of PH with the outcomes of delivery hospitalizations.

## Conclusion

Our study shows that delivery hospitalizations in patients with PH are associated with high risk of various complications. These patients are more likely to be of Black race/ethnicity origin and have other comorbidities. The length of stay is longer compared to patients without PH. Most commonly seen complications include preeclampsia/eclampsia, cardiac arrhythmias, pulmonary edema, and peripartum cardiomyopathy amongst others. PH is most strongly associated with the occurrence of heart failure and pulmonary edema. Prenatal counseling and multidisciplinary care from a cardio-obstetrics team are essential for favorable outcomes in these patients.

## Data Availability

The data related to the manuscript is with the corresponding author and is available on request.

